# SARS-CoV-2, SARS-CoV-1 and MERS-CoV viral load dynamics, duration of viral shedding and infectiousness – a living systematic review and meta-analysis

**DOI:** 10.1101/2020.07.25.20162107

**Authors:** Muge Cevik, Matthew Tate, Ollie Lloyd, Alberto Enrico Maraolo, Jenna Schafers, Antonia Ho

## Abstract

**Background:** Viral load kinetics and the duration of viral shedding are important determinants for disease transmission. We aim i) to characterize viral load dynamics, duration of viral RNA, and viable virus shedding of SARS-CoV-2 in various body fluids and ii) to compare SARS-CoV-2 viral dynamics with SARS-CoV-1 and MERS-CoV.

**Methods:** Medline, EMBASE, Europe PMC, preprint servers and grey literature were searched to retrieve all articles reporting viral dynamics and duration of SARS-CoV-2, SARS-CoV-1 and MERS-CoV shedding. We excluded case reports and case series with < 5 patients, or studies that did not report shedding duration from symptom onset. PROSPERO registration: CRD42020181914.

**Findings:** Seventy-nine studies on SARS-CoV-2, 8 on SARS-CoV-1, and 11 on MERS-CoV were included. Mean SARS-CoV-2 RNA shedding duration in upper respiratory tract, lower respiratory tract, stool and serum were 17.0, 14.6, 17.2 and 16.6 days, respectively. Maximum duration of SARS-CoV-2 RNA shedding reported in URT, LRT, stool and serum were 83, 59, 35 and 60 days, respectively. Pooled mean duration of SARS-CoV-2 RNA shedding was positively associated with age (p=0.002), but not gender (p = 0.277). No study to date has cultured live virus beyond day nine of illness despite persistently high viral loads. SARS-CoV-2 viral load in the upper respiratory tract appears to peak in the first week of illness, while SARS-CoV-1 and MERS-CoV peak later.

**Conclusion:** Although SARS-CoV-2 RNA shedding in respiratory and stool can be prolonged, duration of viable virus is relatively short-lived. Thus, detection of viral RNA cannot be used to infer infectiousness. High SARS-CoV-2 titers are detectable in the first week of illness with an early peak observed at symptom onset to day 5 of illness. This review underscores the importance of early case finding and isolation, as well as public education on the spectrum of illness. However, given potential delays in the isolation of patients, effective containment of SARS-CoV-2 may be challenging even with an early detection and isolation strategy.

**Funding:** No funding was received.

## INTRODUCTION

Viral load kinetics and the duration of viral shedding are important determinants for disease transmission. They determine the duration of infectiousness which is a critical parameter to inform effective control measures and disease modelling. While a number of studies have evaluated SARS-CoV-2 shedding, viral load dynamics and duration of viral shedding reported across studies so far have been heterogenous.^1^ In several case series with serial respiratory sampling, peak viral load was observed just before, or at the time of symptom onset.^2-4^ Viral ribonucleic acid (RNA) shedding was reported to be persistent in the upper respiratory tract and in feces, for over one month after illness onset.^1^ However, the duration of SARS-CoV-2 RNA detection has not been well characterized. A comprehensive understanding of viral load dynamics, length of viral shedding, and how these relate to other factors, such as age and disease severity is lacking.

The aim of this systematic review and meta-analysis was to i) characterize the viral load dynamics of SARS-CoV-2, duration of viral RNA shedding by reverse transcriptase polymerase chain reaction (RT-PCR) and viable virus shedding in various body fluids and ii) compare SARS-CoV-2 viral dynamics with that of SARS-CoV-1 and MERS-CoV.

## METHODS

### Search Strategy

We retrieved all articles reporting viral dynamics and/or the duration of shedding of SARS-CoV-2, SARS-CoV-1 or MERS-CoV in various specimens through systematic searches of major databases including Medline, EMBASE, Europe PMC, pre-print databases (MedRxiv, BioRxiv) and the grey literature from 1 January 2003 to 6^th^ June 2020 using Medical Subject Headings (MeSH) terms (Supplementary Material). We also manually screened the references of included original studies to obtain additional studies. Studies prior to 2003 were excluded since the first recognized case of SARS-CoV-1 was identified in March 2003.

This systematic review was registered in PROSPERO on 29^th^ April 2020 (CRD42020181914) and will be updated in three monthly intervals as a living systematic review.

### Study Selection

Studies were eligible if they met the following inclusion criteria: (1) report on SARS-CoV-2, SARS-CoV-1 or MERS-CoV infection and (2) report viral load kinetics, duration of viral shedding or viable virus. We excluded: (1) review papers; (2) animal studies; (3) studies on environmental sampling; (4) case reports and case series with < 5 participants, due to likely reporting bias; (5) papers where the starting point of viral shedding was not clear or reported from post-discharge and (6) modelling studies with no original data.

### Data Extraction

Two authors (MT and OL) screened and retrieved articles according to the eligibility criteria. Four reviewers (MT, OL, JS, MC) performed full text review and final article selection. From each study, the following variables were extracted as a minimum: name of first author, year of publication, city and country, sample size, median age, sex ratio, time from symptom onset to viral clearance detected by RT-PCR and culture in different specimens, and longest reported time to viral clearance. If these data were not reported, we also contacted the authors to request the data. If available, we extracted data on peak viral load, clinical outcome, and reported factors associated with duration of viral shedding.

#### Risk of bias in included studies

Two authors (OL and JS) independently assessed study quality and risk of bias using the Joanna Briggs Institute (JBI) Critical Appraisal Checklist tools,^5^ which comprise standardized checklists, for the different study designs included in this review. Any disagreements regarding grading of quality were resolved through discussion with a third author (MC).

### Meta-Analysis

For every study included, mean duration of viral shedding and 95% confidence interval (CI) were calculated. The random-effects model (DerSimonian or Laird) was applied to estimate a pooled effect size. Forest plots illustrated the detailed representation of all studies based on the effect size and 95% CI. If not reported, means and standard deviations were derived from sample size, median, interquartile range (IQR), minimum and maximum values.^6^ Heterogeneity between studies were quantified by the I^2^ index and Cochran’s Q test. Publication bias was not assessed as usual appraisal methods are uninformative when meta-analysed studies do not include a test of significance. A weighted meta-regression using an unrestricted maximum likelihood model was performed to assess the impact of potential moderators on the pooled effect size (P-values <0.05 were considered significant). All statistical analyses were performed using Comprehensive Meta-Analysis (CMA) version 3 software (Biostat, Englewood, mNJ).

## RESULTS

The systematic search identified 1486 potentially relevant articles. Three hundred and fifty articles were retrieved for full text review. After reviewing the eligibility criteria, a total of 79 studies on SARS-CoV-2, eight on SARS-CoV-1, and 11 on MERS-CoV were included (Figure 1).

**Figure 1.**
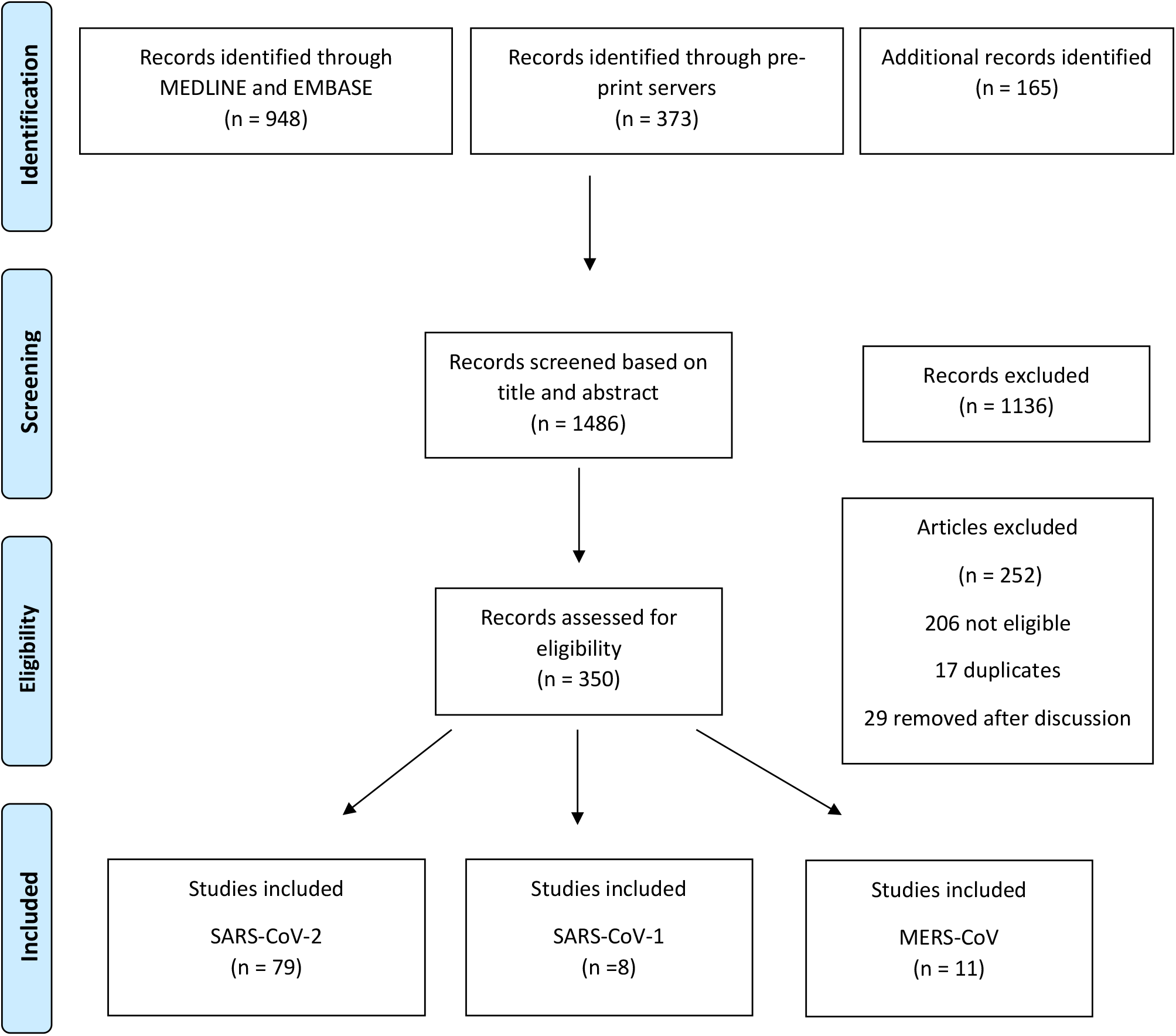
Flowchart describing study selection. *From:* Moher D, Liberati A, Tetzlaff J, Altman DG, The PRISMA Group (2009). *P*referred *R*eporting *I*tems for *S*ystematic Reviews and *M*eta-*A*nalyses: The PRISMA Statement. PLoS Med 6(6): e1000097. doi:10.1371/journal.pmed1000097 For more information, visit www.prisma-statement.org.

### Summary of SARS-CoV-2 studies

Of the 79 papers included, 58 studies were conducted in China (Table 1). Six studies included outpatient or community cases, the remainder comprised hospitalized patients only. Six studies reported viral load dynamics exclusively in children.^7-12^ Two additional studies included children, but data on viral load dynamics were presented in aggregate with adults.^13,14^ One study reported findings in renal transplant patients.^15^

**Table 1:** Summary of included studies.

| Study | Geographical location | Study setting | Study design | Number of patients | Age Median (IQR) | Male sex N (%) | Specimen types |
| --- | --- | --- | --- | --- | --- | --- | --- |
| **SARS-CoV-2** |  |  |  |  |  |  |  |
| **Andersson et al.<sup>62</sup>** | Oxford, UK | Hospital | Case series | 167 | 56 (46-76) | 89 (53) | Serum |
| **Arons et al.<sup>54</sup>** | King's County, USA | Care home | Cross-sectional | 46 | 78.6 ± 9.5* | NR | URT |
| **Bullard et al.<sup>57</sup>** | Manitoba, Canada | Hospital | Case series | 90 | 45 (30-59) | 44 (49) | Respiratory samples (not specified) |
| **Cai et al.<sup>7</sup>** | Shanghai/ Hefei/ Qingdao, China | Hospital | Case series | 10 | 6 | 4 (40) | LRT, blood, stool, urine |
| **Cai et al.<sup>26</sup>** | Shenzhen, China | Hospital | Case series | 298 | 47 (33-61) | 149 (50) | URT |
| **Chang et al.<sup>88</sup>** | Beijing, China | Hospital | Case series | 16 | 35.5 (24-53) | 11 (69) | URT |
| **Chau et al.<sup>53</sup>** | Ho Chi Minh City, Vietnam | Hospital | Case series | 30 | 29 (16-60) | 15 (50) | URT |
| **Chen et al.<sup>27</sup>** | Shanghai, China | Hospital | Case series | 249 | 51 (36-64) | 126 (51) | URT |
| **Chen et al.<sup>89</sup>** | Wuhan, China | Hospital | Case series | 25 | 51.4 ±16.6* | 11 (44) | URT |
| **Chen et al.<sup>28</sup>** | Guangzhou, China | Hospital | Case series | 284 | 48 (33-62) | 131 (46) | URT |
| **Chen et al.<sup>29</sup>** | Wuhan, China | Hospital | Case series | 42 | 51 | 15 (36) | URT, stool, urine |
| **Corman et al.<sup>90</sup>** | Germany | Hospital | Case series | 18 | NR | 12 (67) | Blood |
| **Fan et al.<sup>30</sup>** | Shenyang, China | Hospital | Case series | 55 | 46.8 | 30 (55) | URT, sputum |
| **Fang et al.<sup>31</sup>** | Xiangtan, China | Hospital | Case series | 32 | 41 | 16 (50) | URT, stool, blood |
| **Fu et al.<sup>91</sup>** | Huazhong, China | Hospital | Case series | 50 | 64 (37-87) | 27 (54) | URT |
| **Han et al.<sup>8</sup>** | Chongqing, South Korea | Hospital | Case series | 12 | 6.5 (0.007-16) | 5 (42) | URT, stool |
| **He et al.<sup>16</sup>** | Guangzhou, China | Hospital | Case series | 94 | 46 | 47 (50) | URT |
| **Hu et al.<sup>37</sup>** | Qingdao, China | Hospital | Case series | 59 | 46 (33-57) | 28 (47) | URT |
| **Hu et al.<sup>55</sup>** | Nanjing, China | Hospital | Case series | 24 | 32.5 (21-57) | 8 (33) | URT |
| **Huang et al.<sup>51</sup>** | Guangzhou, China | Hospital | Case series | 27 | NR | 12 (44) | URT |
| **Huang et al.**<sup>23</sup> | Wenzhou, China | Hospital | Case series | 33 | 47 (range 2-84) | 17 (52) | URT, LRT, stool |
| **Huang et al.**<sup>92</sup> | Wuhan, China | Hospital | Retrospective cohort | 200 | 58± 17* | 115 (48) | URT |
| **Hung et al.**<sup>50</sup> | Hong Kong | Hospital | RCT | 127 | 52 (32-62) | 68 (54) | URT, stool |
| **Kim et al.**<sup>4</sup> | Seoul/ Incheon/<br>Seongna, South<br>Korea | Hospital | Case series | 28 | 40 (28-54) | 15 (54) | URT, LRT |
| **Kujawski et al.**<sup>17</sup> | 6 states, USA | Hospital<br>/Outpatient | Case series | 12 | 53 (range 21-68) | 8 (75) | URT, LRT, stool,<br>blood, urine |
| **L'Huillier et al.**<sup>9</sup> | Geneva,<br>Switzerland | Hospital | Case series | 23 | 12 (3.8-14.5) | NR | URT |
| **La Scola et al.**<sup>58</sup> | France | Hospital | Case series | 155 | NR | NR | URT, LRT |
| **Lavezzo et al.**<sup>14</sup> | Vo', Italy | Community | Cross-sectional | Only sample #<br>reported | Mixed | Mixed | URT |
| **Le et al.**<sup>59</sup> | Hanoi, Vietnam | Hospital | Case series | 12 | 29.5* | 3 (25) | URT |
| **Li et al.**<sup>93</sup> | Wuhan China | Hospital | Case series | 36 | 57.5 (52-65) | 23 (64) | URT |
| **Liang et al.**<sup>49</sup> | Wuhan, China | Hospital | Case series | 120 | 61.5 (47-70) | 68 (57) | URT |
| **Ling et al.**<sup>47</sup> | Shanghai, China | Hospital | Case series | 66 | 44 (16-778) | 38 (58) | URT, stool,<br>blood, urine |
| **Liu et al.**<sup>94</sup> | Wuhan, China | Hospital | Case series | 238 | 55 (38.3-65) | 138 (58) | URT |
| **Liu et al.**<sup>32</sup> | Nanchang, China | Hospital | Case series | 76 | 48.3 | 48 (63) | URT |
| **Lo et al.**<sup>95</sup> | Macau, China | Hospital | Case series | 10 | 54 (27-64) | 3 (30) | URT, LRT, stool,<br>urine |
| **Lou B et al.**<sup>96</sup> | Zhejiang, China | Hospital | Case series | 80 | 55 (45-64) | 50 (69) | LRT |
| **Pongpirul et al.**<sup>97</sup> | Bangkok, Thailand | Hospital | Case series | 11 | 61 (28-74) | 6 (55) | URT |
| **Qian et al.**<sup>98</sup> | Ningbo, China | Hospital | Case series | 24 | NR | NR | URT |
| **Quan et al.**<sup>99</sup> | Wuhan/Shenzhen/<br>Xiangyang, China | Hospital | Case series | 23 | 60.3 ±15.3* | 23 (100) | Prostatic<br>secretions all<br>negative (URT) |
| **Sakurai et al.**<sup>43</sup> | Aichi, Japan | Hospital | Case series | 90 | 59.5 (36-68) | 53 (59) | URT |
| **Seah et al.**<sup>100</sup> | Singapore | Hospital | Case series | 17 | NR | NR | Tears |
| **Shastri et al.**<sup>46</sup> | Mumbai, India | Reference lab | Case series | 68 | 37 (range 3-75) | 48 (71) | URT |
| **Shi et al.**<sup>33</sup> | Wuhan, China | Hospital | Case series | 246 | 58 (47-67) | 126 (51) | URT |
| **Song et al.**<sup>101</sup> | Nanjing, China | Hospital | Case series | 13 | 22 – 67 (range only) | 13 (100) | URT, semen, testicular sample |
| **Song et al.**<sup>102</sup> | Beijing, China | Hospital/Outpatient | Case series | 21 | 37 (21-59.5) | 8 (38) | URT |
| **Talmy et al.**<sup>44</sup> | Ramat Gan, Israel | Outpatient | Case series | 119 | 21 (19-25) | 84 (71) | URT |
| **Tan et al.**<sup>34</sup> | Chongqing, China | Hospital | Case series | 142 | NR | NR | URT |
| **Tan et al.**<sup>18</sup> | Chongqing, China | Hospital | Case series | 67 | 49 (10-77) | 35 (52) | URT, LRT, stool, blood, urine |
| **Tan et al.**<sup>10</sup> | Changsha, China | Hospital | Case series | 10 | 7 (1-12) | 3 (30) | URT, stool |
| **Tian et al.**<sup>41</sup> | Beijing, China | Hospital/Outpatient | Case series | 75 | 41.5 (range 0.8 – 88)* | 42 (56) | Respiratory tract sample (not specified further) |
| **To et al.**<sup>19</sup> | Hong Kong, China | Hospital | Case series | 23 | 62 (37-75) | 13 (57) | URT, stool, blood, urine |
| **To et al.**<sup>60</sup> | Hong Kong, China | Hospital | Prospective Cohort | 12 | 62.5 (37-75) | 7 (58) | URT (saliva) |
| **Tu et al.**<sup>103</sup> | Anhui, China | Hospital | Case series | 40 | Viral shedding <10 days:<br>40.86 ± 8.26<br>Viral shedding ≥10 days:<br>45.5 ± 14.60 | 21 (53) | URT |
| **Wang et al.**<sup>104</sup> | Henan, China | Hospital | Case series | 18 | 39 (29-55) | 10 (56) | URT |
| **Wang et al.**<sup>105</sup> | Jinhua, China | Hospital | Case series | 17 | 42 ± 17* | 10 (59) | URT, stool |
| **Wölfel et al.**<sup>20</sup> | Munich, Germany | Hospital | Case series | 9 | NR | NR | URT, blood, urine |
| **Wu et al.**<sup>106</sup> | Hainan, China | Hospital | Case series | 91 | 50 (range 21-83)* | 52 (57) | URT, stool |
| **Wu et al.**<sup>11</sup> | Qingdao, China | Hospital | Case series | 74 | 6 (0.1-15.08 range) | 44 (59) | Stool |
| **Wu et al.**<sup>40</sup> | Zhuhai, China | Hospital | Case series | 74 | 43.8* | 35 (47) | Stool |
| **Wyllie et al.**<sup>21</sup> | New Haven, USA | Hospital | Case series | 44 | 61 (23-92 range)* | 23 (52) | URT (saliva) |
| **Xiao et al.**<sup>45</sup> | Wuhan, China | Hospital | Case series | 56 | 55 (42-68) | 34 (61) | URT |
| **Xiao et al.**<sup>61</sup> | Guangzhou, China | Hospital | Case series | 28 |  |  | Stool |
| **Xu et al.**<sup>38</sup> | Shenzhen/<br>Zhejiang, China | Hospital | Retrospective Cohort | 113 | 52 (42-63) | 66 (58) | URT |
| **Xu et al.**<sup>107</sup> | Shenyang, China | Hospital | Case series | 14 | 48 ± 13.4* | 7 (50) | URT, LRT, serum, conjunctiva |
| **Xu et al.**<sup>12</sup> | Guangzhou, China | Hospital | Case series | 10 | 6.6 | 6 (60) | URT, rectal swab |
| **Yan et al.**<sup>39</sup> | Hubei, China | Hospital | Case series | 120 | 52 (35-63) | 54 (45) | URT |
| **Yang et al.**<sup>56</sup> | Wuhan, China | Hospital | Case series | 78 (45 symptomatic) | Symptomatic: 56 (34-63)<br>Asymptomatic: 37 (26-45) | Symptomatic: 31 (40)<br>Asymptomatic: 11 (33) | URT |
| **Yang et al.**<sup>108</sup> | Shenzhen, China | Hospital | Case series | 213 | 52 (range 2-86) | 108 (51) | URT, LRT |
| **Yongchen et al.**<sup>36</sup> | Nanjing, Xuzhou, China | Hospital | Case series | 21 | 37 | 13 (62) | URT, stool |
| **Young et al.**<sup>22</sup> | Singapore | Hospital | Case series | 18 | 47 | 9 (50) | URT, stool, blood, urine |
| **Zha et al.**<sup>48</sup> | Wuhu, China | Hospital | Case series | 31 | 39 (32-54) | 20 (65) | URT |
| **Zhang et al.**<sup>24</sup> | Beijing, China | Hospital | Case series | 23 | 48 (40-62) | 12 (52) | URT, stool, blood, urine |
| **Zhang et al.**<sup>13</sup> | Shenzhen, China | Hospital | Case series | 56 | Mixed | Mixed | URT, stool |
| **Zheng et al.**<sup>25</sup> | Zhejiang, China | Hospital | Retrospective Cohort | 96 | 53 (33.4-64.8) | NR | LRT, stool, blood, urine |
| **Zhou et al.**<sup>42</sup> | Wuhan, China | Hospital | Case series | 41 | 58 (48-62) | 22 (54) | URT |
| **Zhou et al.**<sup>35</sup> | Wuhan, China | Hospital | Case series | 191 | 56 (46-67) | 119 (62) | URT |
| **Zhou et al.**<sup>52</sup> | Guangzhou, China | Hospital | Case series | 31 | 45 (33-60)<br>37 (28-57) | 4 (44)<br>6 (27) | URT |
| **Zhu et al.**<sup>15</sup> | Wuhan, China | Hospital | Case series | 10 | 49.5 | 8 (80) | URT |
| **Zou et al.<sup>2</sup>** | Zhuhai, China | Hospital/outpatient | Case series | 18 | 59 (range 26-76) | 9 (50) | URT |
| **SARS-CoV-1** |  |  |  |  |  |  |  |
| **Chan et al.<sup>63</sup>** | Hong Kong, China | Hospital | Case series | 415 | 11.3 ± 4.1*<br>37.1 ± 11.2* | 132 (33) | URT, LRT, stool, urine |
| **Chen et al.<sup>64</sup>** | Taiwan | Hospital | Case series | 108 | Stratified | 95 | URT |
| **Cheng et al.<sup>67</sup>** | Hong Kong, China | Hospital | Case series | 1041 | NR | NR | URT, LRT, stool, urine |
| **Kwan et al.<sup>68</sup>** | Hong Kong, China | Hospital | Case series | 12 dialysis<br>33 controls | Dialysis: 58 (range 34-74);*<br>Controls: 57 (range 34-75) | 6 (50) | URT, stools, urine |
| **Liu et al.<sup>65</sup>** | Beijing, China | Hospital | Case series | 56 | 31 (male)<br>34 (female) | 31 (55) | LRT, stool |
| **Leong et al.<sup>69</sup>** | Singapore | Hospital | Case series | 64 | 35.2 (17-63 range)* | 16 (25) | URT, stool, blood, urine |
| **Peiris et al.<sup>70</sup>** | Hong Kong, China | Hospital | Case series | 75 | 39.8 (SD 12.2) | 0.92 | URT |
| **Xu et al.<sup>109</sup>** | Beijing, China | Hospital | Case series | 54 | NR | NR | LRT, blood, urine |
| **MERS-CoV** |  |  |  |  |  |  |  |
| **Al Hosani et al.<sup>73</sup>** | Abu Dhabi, UAE | Hospital/community | Case series | 65 | 20 -59 | 43 (66) | LRT |
| **Al-Jasser et al.<sup>110</sup>** | Riyadh, Saudi Arabia | Hospital | Case series | 167 | 46.71* | 142 (57) | URT |
| **Alkendi et al.<sup>111</sup>** | Tawam/Al Ain, UAE | Hospital | Case series | 58 | 43.5 | 41 (71) | URT |
| **Arabi et al.<sup>75</sup>** | Saudi Arabia | Hospital | Cohort | 330 | 58 (44-69) | 225 (68) | URT |
| **Corman et al.<sup>77</sup>** | Riyadh, Saudi Arabia | Hospital | Case series | 37 | 69 (24-90)* | 27 (39) | URT, LRT, stool, blood, urine |
| **Hong et al.<sup>76</sup>** | Seoul, South Korea | Hospital | Case series | 30 | 49* | 19 (63) | Blood |
| **Min et al.<sup>71</sup>** | Seoul/others, South Korea | Hospital | Case series | 14 | 62 | 6 (35) | LRT, serum |
| **Muth et al.<sup>78</sup>** | Riyadh, Saudi Arabia | Hospital | Case series | 32 | 66 (24-90) | 24 (75) | LRT |
| **Oh et al.<sup>74</sup>** | Seoul, South Korea | Hospital | Case series | 17 | NR | NR | URT, LRT, serum |
| **Park et al.<sup>112</sup>** | Seoul, South Korea | Hospital | Case series | 17 | NR | NR | URT, LRT |
| **Shalhoub et al.<sup>72</sup>** | Jeddah, Saudi Arabia | Hospital | Retrospective cohort | 32 | 65 | 14 (44) | LRT, serum |

Abbreviations: UK, United Kingdom, USA; United States of America; UAE, United Arab Emirates; RCT, randomised controlled trial; URT, upper respiratory tract; LRT, lower respiratory tract; NR, not reported.
\* Mean ± standard deviation (or range if stated).

### Median duration of viral shedding

In total, 61 studies reported median or maximum viral RNA shedding in at least one body fluid and six studies provided duration of shedding stratified by illness severity only. Of those, 43 (3229 individuals) reported duration of shedding in upper respiratory tract (URT), seven (260 individuals) in lower respiratory tract (LRT), 13 (586 individuals) in stool, and 2 studies (108 individuals) in serum samples were eligible for quantitative analysis. Means viral shedding durations were 17.0 days (95% CI, 15.5-18.6), 14.6 days (95% CI, 9.3-20.0), 17.2 days (95% CI, 14.4-20.1) and 16.6 days (95% CI, 3.6-29.7), respectively (Figures 2 to 5). Maximum duration of RNA shedding reported in URT, LRT, stool and serum were 83, 59, 35 and 60 days, respectively.

**Figure 2:**
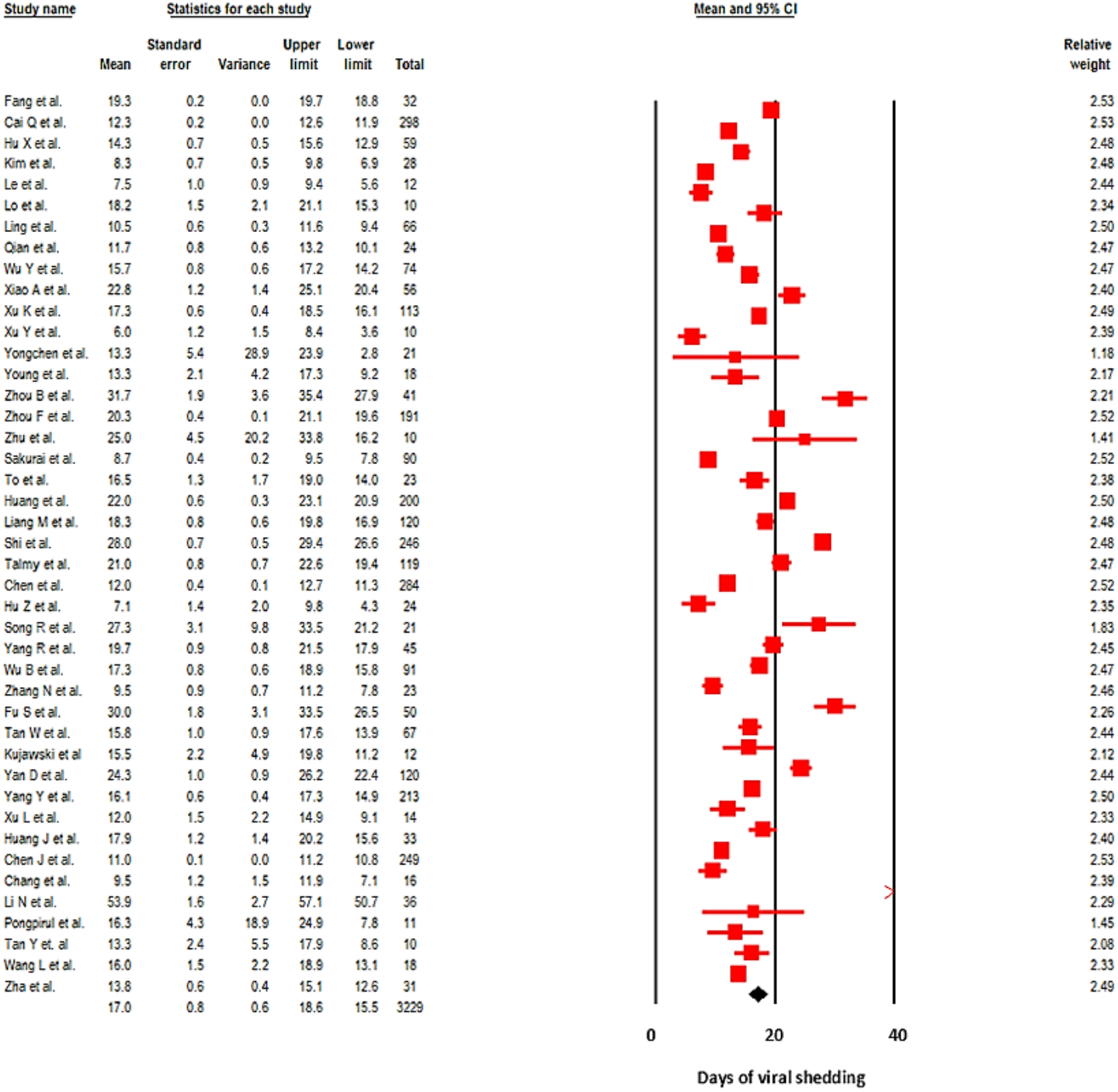
Pooled mean duration (days) of SARS-CoV-2 shedding from the upper respiratory tract (random-effects model). Note: the overall effect is plotted as a black square. Test for heterogeneity: Q-value = 4076,08, df(Q) = 42, p < 0.001, I^2^ = 99%.

**Figure 3:**
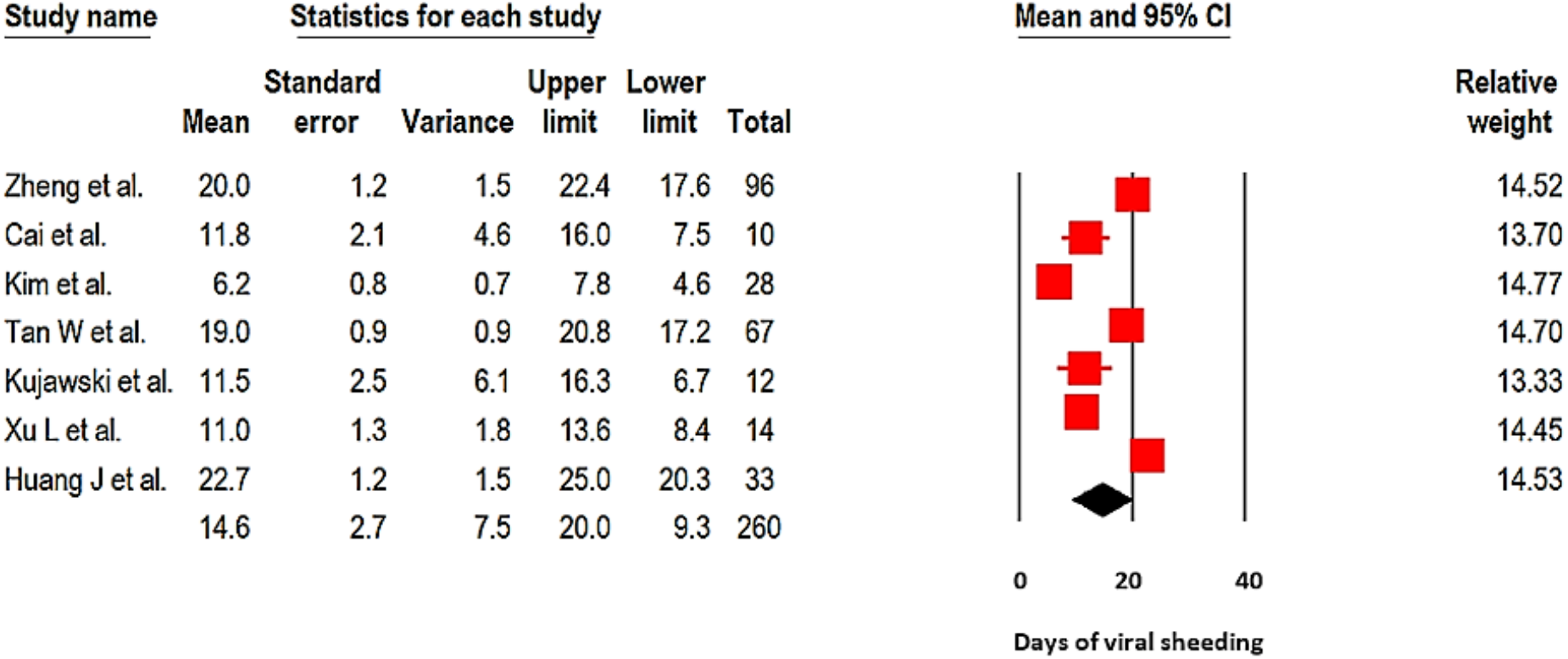
Pooled mean duration (days) of SARS-CoV-2 shedding from the lower respiratory tract (random-effects model). Note: the overall effect is plotted as a black square. Test for heterogeneity: Q-value = 203.3, df(Q) = 6, p < 0.001, I^2^ = 97%.

**Figure 4.**
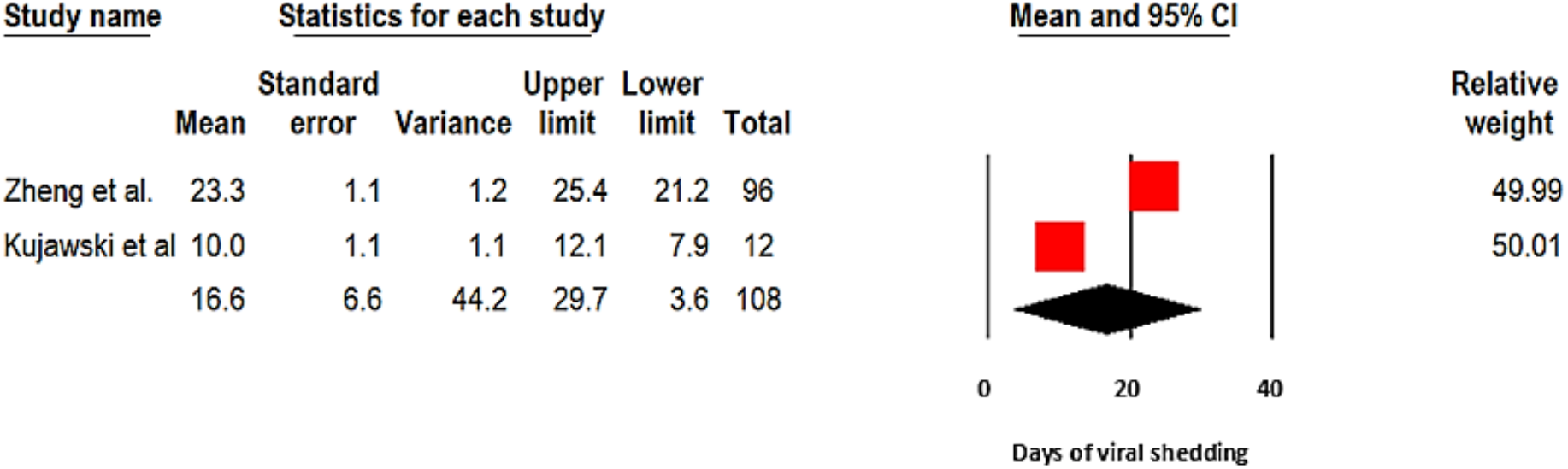
Pooled mean duration (days) of SARS-CoV-2 shedding in the blood (random-effects model). Note: the overall effect is plotted as a black square. Test for heterogeneity: Q-value = 77,6, df(Q) = 1, p < 0.001, I^2^ = 99%.

**Figure 5.**
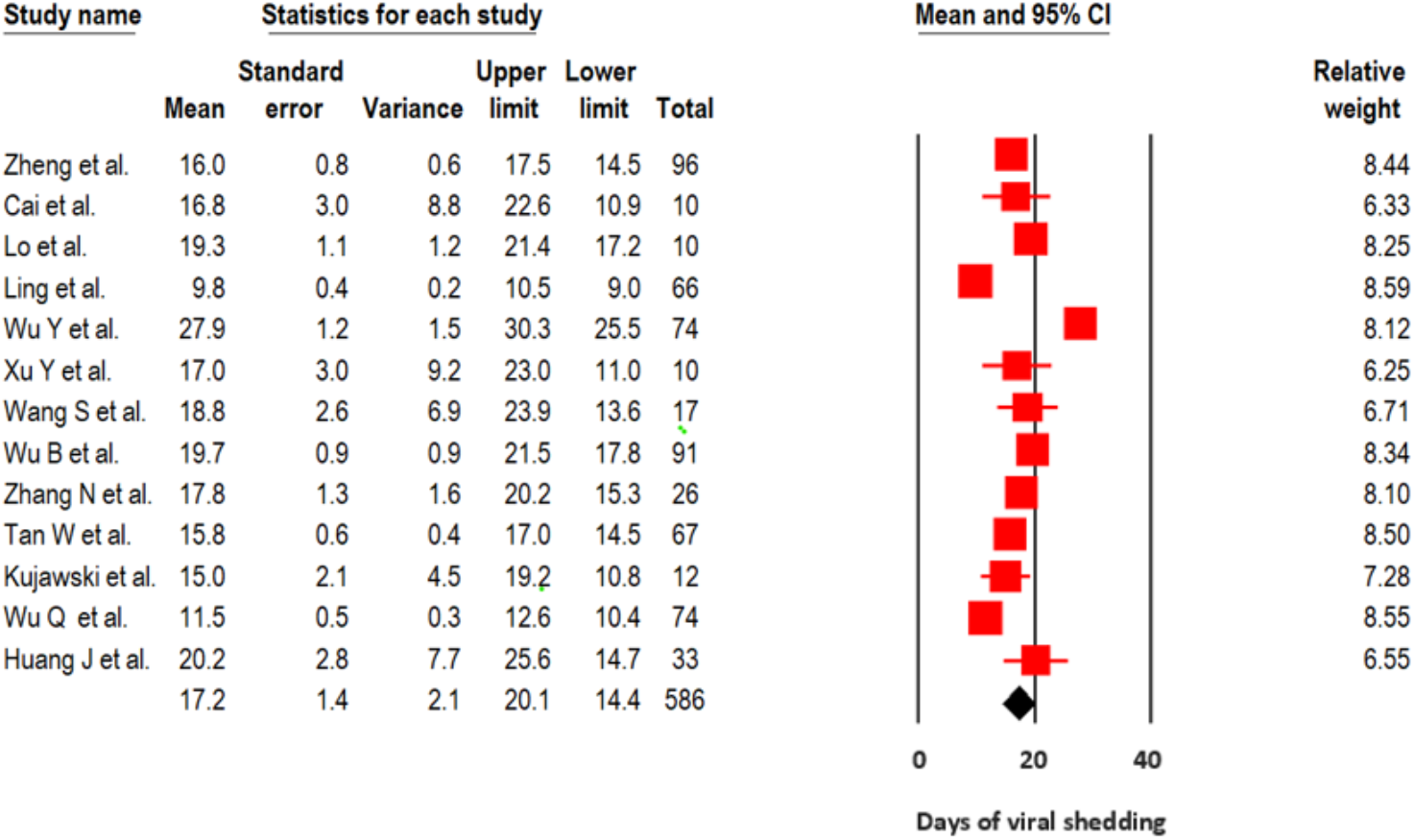
Pooled mean duration (days) of SARS-CoV-2 shedding from the stool (random-effects model). Note: the overall effect is plotted as a black square. Test for heterogeneity: Q-value = 356.0, df(Q) = 12, p < 0.001, I2 = 96.6%.

Studies reporting duration of viral shedding in URT and stool samples were eligible for meta-regression analysis. Pooled mean viral shedding duration was positively associated with age (slope: +0.304; 95% CI, +0.115 to +0.493; p = 0.002 Fig 6), but not gender (p = 0.277, Supplementary Fig 3). When adjusted for the proportion of male subjects in a multivariable analysis, mean age was positively associated with the mean duration of viral shedding in URT specimens (p = 0.003). There was a positive but non-significant association between mean age and duration of shedding in stool (p = 0.37) (Supplementary Figure 4).

**Figure 6.**
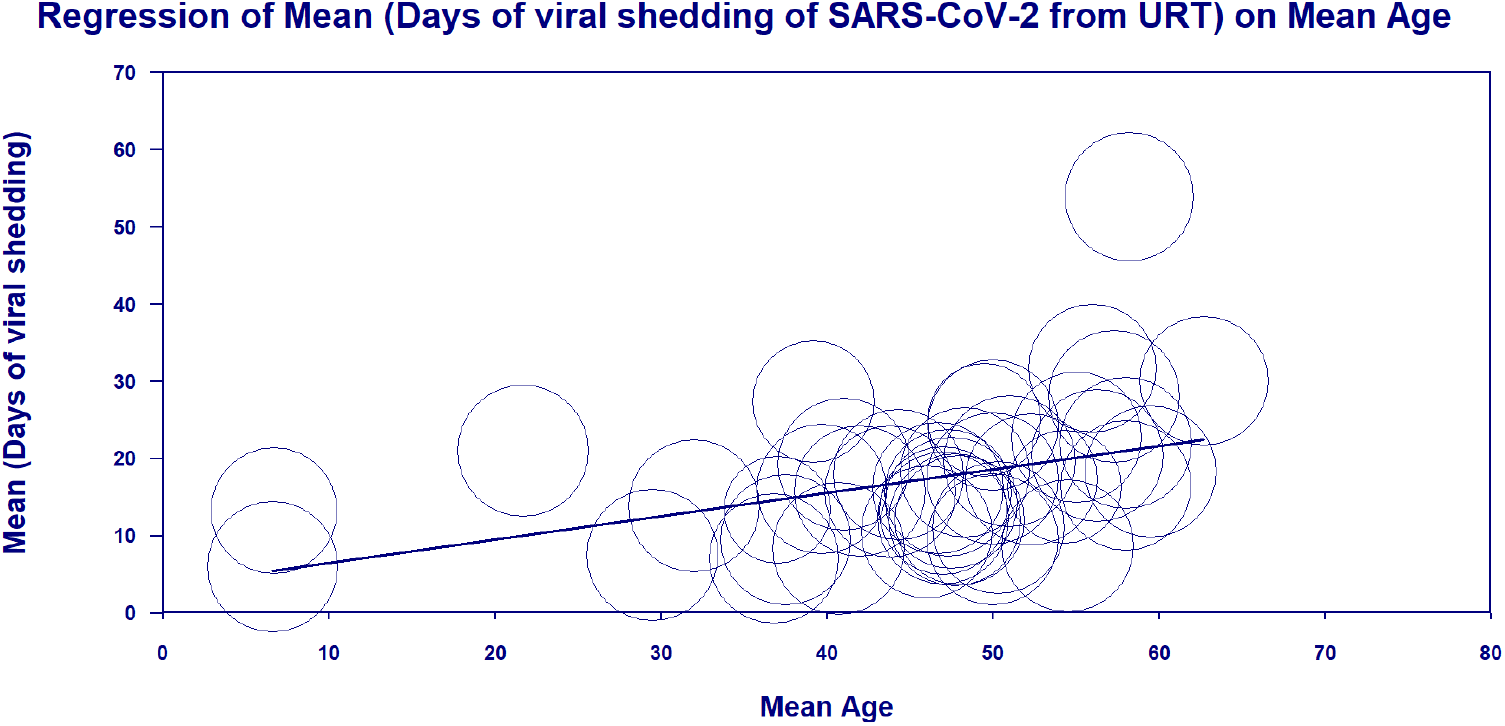
Meta-regression bubble plot of the impact of age on mean SARS-CoV-2 shedding from the upper respiratory tract. URT: upper respiratory tract. Note: the plot was built upon 41 studies (no data on mean age from the study of Qian et al.^98^). A random-effects model was used.

### Peak viral load

The majority of studies evaluating SARS-CoV-2 viral load in serial URT samples demonstrated peak viral loads within the first week of symptom onset. ^2,4,8,16-24^ The highest viral loads were reported either soon after or at the time of symptom onset^2,8,16,23,24^ or at day 3-5 of illness^4,18,20,22^ followed by a consistent decline.

Five studies that evaluated viral load dynamics in LRT samples observed a peak viral load in the second week of illness.^4,18,20,23,25^ In contrast, the dynamics of SARS-CoV-2 shedding in stool is more erratic, with highest viral loads reported on day 7,^18^ 2-3 weeks,^24,25^ and up to 5-6 weeks after symptom onset.^23^ While several studies reported significantly higher viral titres in stool compared to respiratory samples,^8,25^ Huang *et al*. reported lower viral load in stool than in both LRT and URT samples early in the disease course.^23^

### Severity and association with duration of viral shedding

In total, 20 studies evaluated duration of viral RNA shedding based on disease severity. The majority (n=13) reported longer duration of viral shedding in patients with severe illness than those with non-severe illness, ^18,25-36^ while five studies reported similar shedding durations according to disease severity in URT samples^17,19,37-39^ and one study in stool samples.^40^ Only one study reported shorter viral shedding in moderate to severe illness compared to mild to moderate illness.^41^ Six studies have performed comparative analysis based on severity of illness;^18,25,27,28,38,39^ the majority (n=5) demonstrated significantly longer duration of shedding among the severe illness group compared to the non-severe patients and only one study observed no difference.^39^ (Table 2).

**Table 2:** Severity of illness and viral dynamics.

| Study | Classification of severity | Median duration - days (IQR) | Viral dynamics in severe patients compared to non-severe patients | P-value |
| --- | --- | --- | --- | --- |
| **Chen *et al.*<sup>27</sup>** | ICU vs. non-ICU patients | 11 | Median time to viral clearance significantly longer in ICU vs. non-ICU patients (HR=3.17, 95% CI, 2.29-4.37) | Only HR provided |
| **Chen *et al.*<sup>28</sup>** | China CDC guideline (version 7) | 12 (8-16) | Shedding duration varies by severity: asymptomatic 6 days; mild 10 days; moderate 12 days; serious 14 days; critical 32 days | <0.0001 |
| **Tan *et al.*<sup>18</sup>** | China CDC guideline (version 6) | NP: 12<br>Any sample: 22 | Viral shedding significantly longer in severe patients: any sample 23 vs. 20 days (note NP: 14 vs. 11 days – non-significant) | p=0.023 (any sample) |
| **Xu *et al.*<sup>38</sup>** | WHO criteria | 17 (13-32) | Higher proportion of severe patients had shedding >21 days (34.2% vs. 16.2%) | 0.49 |
| **Yan *et al.*<sup>39</sup>** | China CDC guideline (version 6) | 23 (18-32) | No difference in shedding duration (general 23 days vs. severe 26 days vs. critical 28 days) | 0.51 |
| **Zheng *et al.*<sup>25</sup>** | China CDC guideline (version 6) | Resp: 18 (13-29) | Shedding duration significantly longer in severe patients (21 vs 14 days) in respiratory samples.<br>No difference in shedding duration in stool/serum | p=0.04 |

Abbreviations: IQR, interquartile range; ICU, intensive care unit; HR, hazard ratio; CDC, Centers for Disease Control and Prevention; WHO, World Health Organization.

### Other factors associated with prolonged shedding

All but one study^42^ (n=10) that examined the impact of age on SARS-CoV-2 shedding identified an association between older age and prolonged viral RNA shedding.^25,26,28,33,37-39,43-45^ Three studies identified age as an independent risk factor for delayed viral clearance.^25,26,38^ Male sex was also associated with prolonged shedding,^25,38,46^ and the association remained significant even when patients were stratified based on illness severity.^25,38^ Corticosteroid treatment was associated with delayed viral clearance in four studies,^33,38,47,48^ and one study that recruited 120 critically ill patients, found no difference between corticosteroid and control groups.^49^

In a phase 2 open-label study evaluating interferon beta-1b, lopinavir–ritonavir, and ribavirin a shorter duration of viral shedding was seen with combination treatment compared to the control.^50^ None of the antiviral regimens (chloroquine, oseltamivir, arbidol, and lopinavir/ritonavir) independently improved viral RNA clearance.^28,51^ In a retrospective study of 284 patients, lopinavir/ritonavir use was associated with delayed viral clearance even after adjusting for confounders.^28^

### Asymptomatic SARS-CoV-2 shedding

Twelve studies reported on viral load dynamics and/or duration of viral shedding among patients with asymptomatic SARS-CoV-2 infection (Table 3); two demonstrated lower viral loads among asymptomatic patients compared to symptomatic patients,^8,52^ while four studies found similar initial viral loads. ^13,14,53,54^ However, Chau *et al* reported significantly lower viral load in asymptomatic patients during the follow up compared to symptomatic patients.^53^ Faster viral clearance was observed in asymptomatic individuals in five out of six studies.^13,28,53,55,56^ The exception Yongchen *et al*., found longer shedding duration among asymptomatic cases, but the difference was not significant.^36^

**Table 3:** SARS-CoV-2 viral dynamics in asymptomatic patients compared to symptomatic patients.

|  | Median duration – days (IQR) | Viral dynamics in asymptomatic patients compared to symptomatic patients | P-value |
| --- | --- | --- | --- |
| **Arons *et al.*<sup>54</sup>** | NR | No difference in viral load | NS |
| **Chau *et al.*<sup>53</sup>** | NR | Initial viral load similar. Asymptomatic patients had significantly lower viral load during the follow up compared to symptomatic patients and faster viral clearance in asymptomatic, compared to symptomatic individuals | 0.027 |
| **Chen *et al.*<sup>28</sup>** | 6 (3.5-10) | Significantly shorter duration of viral shedding among asymptomatic cases (median 6 days, IQR 3.5-10), with increasing shedding duration associated with increasing illness severity | <0.0001 |
| **Han *et al.*<sup>8</sup>** | NR | Symptomatic children had higher initial RNA load in nasopharyngeal swab specimens than asymptomatic children (9.01 vs. 6.32 log<sub>10</sub> copies/mL; p = 0.048). | 0.048 |
| **Hu *et al.*<sup>55</sup>** | 6 (2-12) | Asymptomatic patients had shorter duration of viral shedding compared to pre-symptomatic patients (median duration of SARS-CoV-2 positivity was 6.0 (2.0 - 12.0) compared to 12.0 (12.0 - 14.0)) | NR |
| **Lavezzo *et al.*<sup>14</sup>** | NR | No difference in viral load | NS |
| **Le *et al.*<sup>59</sup>** | 9 | NR | N/A |
| **Sakurai *et al.*<sup>43</sup>** | 9 (6-11) | NR | N/A |
| **Yang *et al.*<sup>56</sup>** | 8 (3-12) | Significantly shorter duration of viral shedding from nasopharynx swabs was observed among asymptomatic compared to symptomatic patients | P= .001 |
| **Yongchen *et al.*<sup>36</sup>** | 18 (5-28) | Longer shedding duration among asymptomatic cases (median 18 days, range 5-28), compared to non-severe (10 days, range 2-21) and severe (14 days, range 9-33) cases | NS |
| **Zhang *et al.*<sup>13</sup>** | 9.63 | Initial viral load similar, viral clearance occurred earlier in the asymptomatic (9.6 days) and symptomatic individuals (9.7 days, compared to pre-symptomatic group (13.6 days) |  |
| **Zhou *et al.*<sup>52</sup>** | NR | Significantly higher viral load in symptomatic (n=22) compared to asymptomatic (n=9) patients (median cycle threshold (Ct) value 34.5 vs. 39.0, respectively) but duration of shedding was similar |  |

Abbreviations: IQR, interquartile range; RNA, ribonucleic acid; NR, not reported; NS, non-significant; N/A, not applicable

### Live virus detection

We identified 11 studies that attempted to isolate live virus. All eight studies that attempted virus isolation in respiratory samples successfully cultured viable virus within the first week of illness, ^9,17,20,54,57-60^ No live virus was isolated from any respiratory samples taken after day 8 of symptoms in three studies,^20,57,58^ or beyond day 9 in two studies^17,54^ despite persistently high viral RNA loads. One study demonstrated the highest probability of positive culture on day 3 of symptoms.^57^ Arons *et al*. cultured viable virus 6 days before typical symptom onset, however onset of symptom was unclear.^54^

The success of viral isolation correlated with viral load quantified by RT-PCR. No successful viral culture was obtained from samples with a viral load below 10^6^ copies/ml, ^20^ Ct values >24,^57^ or >34,^54,58^ with culture positivity declining with increasing Ct values.^58^ Several other studies cultured live virus from RT-PCR positive specimens; however, they did not correlate these results with viral load titres.^9,59,60^

Only one study reported the duration of viable virus shedding in respiratory samples; the median time to clearance from URT and LRT samples was 3.5 and 6 days, respectively.^20^ Arons *et al*. cultured viable virus in one out of three asymptomatic cases from the respiratory tract.^54^

Viral culture was successful in two of three RNA-positive patients in one study, but the time points from symptom onset were not reported.^61^ Andersson *et al*. failed to culture virus from 27 RT-PCR positive serum samples.^62^

### Summary of SARS-CoV-1 and MERS studies

Eight studies on SARS-CoV-1 were included; the majority of studies did not report mean or median duration of viral shedding thus, were not eligible for quantitative analysis. The maximum duration of viral shedding reported was 8 weeks in URT,^63,64^ 52 days in LRT,^63,65^ 6-7 weeks in serum,^66^ and 126 days in stool samples.^63,65,67-69^ Dialysis patients had longer viral shedding in stool compared to non-dialysis patients.^68^ Studies that have evaluated SARS-CoV-1 kinetics found low viral load in the initial days of illness, increasing after the first week of illness in URT samples, peaking at day 10,^70^ or day 12-14,^67^ and declining after week 3-4.^64^ High viral loads correlated with severity of illness^64^ and poor survival.^64^ While Chen *et al*. identified an association between younger age and lower viral titers, ^64^ Leong *et al*. found no difference.^69^ Viable SARS-CoV-1 was isolated from stool and respiratory samples up to 4 weeks, and urine specimens up to day 36.^63,66^ All attempts to isolate virus from RT-PCR–positive stool specimens collected >6weeks after disease onset failed.^65^ The isolation probability for stool samples was approximately 5 to 10 times lower compared to respiratory specimens.^63^

We identified 11 studies on MERS-CoV. Three studies (324 subjects) reporting MERS-CoV shedding in URT and four studies (93 subjects) in LRT were included in the quantitative analysis. The mean shedding duration was 15.3 days (95% CI, 11.6 – 19.0) and 16.6 days (95% CI, 14.8 – 18.4), respectively (Supplementary Figures 1 and 2). Only one study reported duration of viral shedding in serum with a median of 14 days and max of 38 days.^71^ In a small study, mortality rates were higher in patients with viraemia.^72^ In URT and LRT specimens, prolonged shedding was associated with illness severity^73,74^ and survival^75^ with the shortest duration observed in asymptomatic patients.^73^ Peak viral loads were observed between days 7 to 10 and higher viral loads was observed among patients with severe illness and fatal outcome.^71,73,74,76,77^ Differences in viral loads between survivors and fatal cases was more pronounced in the second week of illness (P< 0.0006).^77^ The proportion of successful viable culture was 6% in respiratory samples with a viral load values below 10^7^copies/ml.^78^

### Qualitative analysis

All but 11 studies (6 cohort studies, 2 cross-sectional studies, and 1 RCT on SARS-CoV-2 and 2 cohort studies on MERS-CoV) were case series, the majority of which recruited non-consecutive patients and therefore prone to possible selection bias. (Supplementary Table 1)

## DISCUSSION

This systematic review and meta-analysis provide comprehensive data on the viral dynamics of SARS-CoV-2 including the duration of RNA shedding and viable virus isolation. Our findings suggest that while patients with SARS-CoV-2 infection may have prolonged RNA shedding, median time to live virus clearance from upper and lower respiratory tract samples were 3.5 days and 6 days, respectively. No live virus isolated from culture beyond day nine of symptoms despite persistently high viral RNA loads, thus emphasizing that the infectious period cannot be inferred from the duration of viral RNA detection. This finding is supported by several studies demonstrating a relationship between viral load and viability of virus, with no successful culture from samples below a certain viral load threshold.

SARS-CoV-2 viral load appears to peak in the URT within the first week of illness, and later in the LRT. In contrast, peaks in SARS-CoV-1 and MERS-CoV viral loads in the URT occurred at days 10-14 and 7-10 days of illness, respectively. Combined with isolation of viable virus in respiratory samples primarily within the first week of illness, patients with SARS-CoV-2 infection are likely to be most infectious in the first week of illness. Several studies report viral load peaks during the prodromal phase of illness or at the time of symptom onset,^2,4,8,16-23^ providing a rationale for the efficient spread of SARS-CoV-2. This is supported by the observation in contact tracing studies that the highest risk of transmission occurs during the prodromal phase or early in the disease course.^79,80^ No secondary cases were identified beyond 5 days after the symptom onset.^81^ Although modelling studies estimated potential viral load peak before symptom onset, we did not identify any study that confirms pre-symptomatic viral load peak.^16^

Emerging evidence suggests a correlation between virus persistence and disease severity and outcome.^18,25,27-29,38^ This is consistent with the viral load dynamics of influenza, MERS-CoV, and SARS-CoV-1 whereby severe disease was also associated with prolonged viral shedding.^73,74,82^ However, more studies are needed to understand the duration of viable virus in patients with severe illness.

Similar to SARS-CoV-1, SARS-CoV-2 can be detected in stool for prolonged periods, with high viral loads detected even after 3 weeks of illness. A clear difference between SARS-CoV-1 and MERS-CoV is the detection of viral RNA in stool. In SARS-CoV-1, RNA prevalence in stool samples was high, with almost all studies reporting shedding in stool. Although viable SARS-CoV-1 was isolated up to 4 weeks of illness, fecal-oral transmission was not considered to be a primary driver of infection. Whereas in MERS-CoV, none of the studies reported duration of viral shedding in stool and RNA detection was low.^77,83^ To date, only a few studies have demonstrated viable SARS-CoV-2 in stool.^61,84^ Thus, the role of fecal shedding in viral transmission remains unclear.

Viral loads at the start of infection appear to be comparable between asymptomatic and symptomatic patients infected with SARS-CoV-2. Nevertheless, most studies demonstrate faster viral clearance among asymptomatic individuals. This suggests similar transmission potential among both groups at the onset of infection, but a shorter period of infectiousness in asymptomatic patients. This is in keeping with viral kinetics observed with other respiratory viruses such as influenza and MERS-CoV, in which people with asymptomatic infection have a shorter duration of viral shedding than symptomatic individuals.^73,85^ However, there are limited data on the shedding of infectious virus in asymptomatic individuals to quantify their transmission potential to inform policy on quarantine duration in the absence of testing.

This is the first study that has comprehensively examined and compared SARS-CoV-2, SARS-CoV-1 and MERS-CoV viral dynamics and performed a meta-analysis of viral shedding duration. Our study has limitations. First, some patients in the included studies received a range of treatments, including steroids and antivirals, which may have modified the shedding dynamics. Second, most of the included studies are case series, which are particularly vulnerable to selection bias. Third, our meta-analysis identified substantial study heterogeneity, likely due to differences in study population, follow up and management approaches. Further, shedding duration is reported as median ± IQR for most studies, but meta-analysis necessitates conversion to mean ± SD.^6^ The validity of this conversion is based on the assumption that duration of viral shedding is normally distributed, which may not apply to some studies. Lastly, although there is likely a broad overlap, the true clinical window of infectious shedding may not entirely align with viral culture duration.

We identified a systematic review of SARS CoV-2 viral load kinetics that included studies published up until 12 May 2020.^86^ This review included many studies that did not meet our eligibility criteria, including 26 case reports and 13 case series involving <5 individuals; these are prone to significant selection bias, reporting atypical cases with prolonged viral shedding. Additionally, the review included studies that reported viral shedding duration from the time of hospital admission or initial PCR positivity, rather than symptom onset. Furthermore, no meta-analysis of the duration of viral shedding was performed.

This review provides detailed understanding about the available evidence to date on viral dynamics of SARS-CoV-2 and has implications for pandemic control strategies and infection control practices. Although SARS-CoV-2 RNA shedding can be prolonged in respiratory and stool samples, the duration of viable virus is short-lived, with culture success associated with viral load levels. No study has reported live SARS-CoV-2 beyond day nine to date. Most studies detected the SARS-CoV-2 viral load peak within the first week of illness. These findings highlight that isolation practices should be commenced with the start of first symptoms including mild and atypical symptoms that precede more typical COVID-19 symptoms. This systematic review underscores the importance of early case finding and isolation, as well as public education on the spectrum of illness. However, given potential delays in the isolation of patients, effective containment of SARS-CoV-2 may be challenging even with an early detection and isolation strategy.^87^

## Data Availability

All data fully available

## Authors contributions

M. Cevik: conceptualisation, methodology, investigation, data curation, writing – original draft. M. Tate: investigation, data curation, writing – original draft; O Lloyd: investigation, data curation, writing – review and editing; A. E. Maraolo: formal analysis, writing – original draft; J. Schafers: investigation, data curation, writing – review and editing; A Ho: conceptualisation, methodology, data curation, writing – original draft, supervision.

## Acknowledgements

We would like to thank Vicki Cormie at the University of St Andrews for assistance with the search and obtaining papers not readily accessible.

